# Factors associated with the incidence rate of HIV viral rebound among children and adults receiving antiretroviral therapy in Malawi using the Laboratory Management information System: 2011-2020

**DOI:** 10.1101/2021.10.07.21264672

**Authors:** Wingston Ng’ambi, Janne Estill, Andreas Jahn, Erol Orel, Tiwonge Chimpandule, Rose Nyirenda, Olivia Keiser

## Abstract

**Objective:** To describe the trends in the incidence rate of HIV VL rebound and assess the factors associated with the incidence rate of HIV VL rebound among the persons receiving ART in Malawi between 2011 and 2020.

**Methods:** We analyzed routinely collected patient-level HIV VL data extracted from the Malawi Laboratory Management Information System (LIMS). We fitted a multivariable Cox regression model of having HIV VL rebound (>1000 copies/mL) using a forward step-wise selection method, with age and sex entered as a priori variables. We presented both crude and adjusted hazard ratios (aHR) of HIV VL rebound for the various independent variables. Statistical significance was set at P<0.05.

**Results:** We evaluated 230,465 adults and children receiving ART and having at least one viral load test between 2011 and 2020. The median time between ART initiation and having a VL sample was 55 months (interquartile range: 25-92). The aHR for having HIV VL rebound were: 0.79 for females (95%CI:0.77-0.80, P<0.001) compared to males, 0.95 (95%CI: 0.94-0.98, P<0.001) for patients from rural compared to urban areas and 0.57 (95%CI:0.56-0.58, P<0.001) for routine compared to targeted VL tests. There was a decreasing trend in the aHR of patients having HIV VL rebound with increasing age.

**Conclusion:** This is the first national analysis of Malawi HIV VL data from LIMS. Our findings show the need to take into account the geographic and demographic characteristics of the patients in order to persistently suppress HIV VL and consequently achieve 95% HIV VL suppression by 2030.

## INTRODUCTION

The World Health Organization (WHO) has recommended immediate initiation of antiretroviral therapy (ART) for all people living with HIV since 2015 although the WHO still recommends measuring the CD4 cell counts at enrolment into HIV care [1] [2]. The CD4 cell count is critical in guiding the prophylactic and diagnostic interventions, for example prioritizing testing for opportunistic infections such as Cryptococcus or tuberculosis [3]. The main benefit of ART is the suppression of HIV□1 viral replication [2] [4]. The benefit of suppression of viral load (VL) (defined by WHO as ≤1000 HIV□RNA copies/mL) is in the reduction of morbidity and mortality among patients living with HIV and onward transmission of the virus [1]. Since 2013, WHO has recommended VL monitoring approach to detect treatment failure following ART initiation [1] [2] [3]. WHO guidelines were further refined in 2016 and currently recommend routine VL testing is done yearly thereafter among patients stable on ART [2]. Patients on regimens non-nucleoside reverse transcriptase inhibitor (NNRTI) based regimens with two consecutive unsuppressed VL measurements (>1000 copies/mL) despite enhanced adherence counselling, taken within a three□month interval and at least six months after starting ART are considered to have treatment failure [1]. However, patients on Integrase Strand Transfer Inhibitor (INSTI) or Protease inhibitors (PIs) based regimens need a genotype after high FUP VL to distinguish treatment failure due to drug resistance from poor adherence which is much more common as an underlying reason on these regimens. Treatment failure indicates either some degree of drug resistance, or that ART has not been taken properly [1].

Malawi achieved two out of three the 2020 Joint United Nations Programme on HIV/AIDS (UNAIDS) 90-90-90 target with 91% of the people living with HIV being aware of their status, 83% of those aware of their status being on ART, and 90% of those on ART being virally suppressed in 2020 [5]. Over the past decade, there has been an increasing trend in the number of persons started on ART in Sub-Saharan African (SSA) settings [6]. Therefore, it is important to sustain treatment success and limit the development of treatment failure [6]. Despite the WHO recommending routine uptake of VL monitoring since 2013, the uptake has been sub-optimal especially in SSA due to limited testing capacity or shortage of staff to conduct the testing [7]. The uptake of VL testing has varied across countries, age, antiretroviral drugs and sex. For example, the proportion of patients with at least one VL test by mid-2016 was 19% in Malawi, 11% in Cote d’Ivoire, 49% in Kenya, 43% in Namibia, 9% in Tanzania and 22% in Uganda. [2] [7].

The Malawi Ministry of Health (MoH) has been implementing routine VL monitoring since 2012. Before 2018, patients on ART were supposed to have an HIV VL every two years with the first one to be taken at six months after start of ART treatment (i.e., 6, 24, 48, 72, etc. for routine HIV VL assessment). Since 2018, the ART patients are supposed to have a routine HIV VL test every year. The Malawi MoH therefore tracks the prevalence and incidence of VL suppression amongst the persons who are receiving ART. Since the introduction of this program no in-depth analyses have been done to assess the trends in incidence of HIV VL rebound amongst the persons receiving ART at national level in Malawi. However, in-depth analyses are necessary for a greater understanding of HIV VL monitoring and its quality improvement as well as ensuring the achievement of the UNAIDS target of ending HIV/AIDS by 2030. This study therefore aims to describe the trends in the incidence rate of VL failure, and assess the factors associated with temporal trends in the incidence rate of VL failure among persons receiving ART in Malawi between 2011 and 2020.

## METHODS

### Study design

We used patient-level HIV VL data from the Laboratory Information Management Systems (LIMS) national database containing data collected between 2012 and 2020 in Malawi [8]. The LIMS database is a central data repository for all molecular laboratory tests in Malawi managed by the Diagnostics Department of the Ministry of Health. The LIMS database contains individual level HIV VL data from all the districts and ART facilities in Malawi. The database has inbuilt tools for performing data quality assessment like range checks and other associated validation rules.

### Data collection

The source of patient level characteristics is the lab requisition forms that are filled by staff collecting the samples, usually the HIV Diagnostic Assistants (HDAs). This can be a considerable challenge as the HDAs are often not fully aware of to the details of the regimen and other patient characteristics. Furthermore, the data are captured as a single entry by clerks at the lab receptions, which explains some of the gaps and obvious inconsistencies in the data.

The LIMS dataset has no reliable unique patient identifier, which makes it hard to identify sequential samples from the same patients. We were able to re-identify some patients on the basis of their characteristics from LIMS. This makes it obvious that many samples are mislabeled as “routine scheduled” when we can easily deduce that the interval from the last sample is too short for the scheduled monitoring milestones. Such samples are re-classified as follow-up. The routine HIV VL are currently being done yearly for a patient that has been on ART for at least 12 months. However, when a clinician has suspicion that the patient may be failing on ART then a targeted VL is done based on need.

### Data Management

The data were managed in Stata v16.0 (Stata Corp., Texas, USA). All HIV VL data collected between 1 January 2012 and 31 December 2020 were included in this study. The response variable was unsuppressed VL (defined as having a VL of at least 1000 copies [9]. The independent variables were: age at sample collection, year of sample collection, sex, facility location (rural/urban), reason for VL testing (routine/targeted), current regimen, sample type (plasma/dry blood spot (DBS)) and region (north/center/south). The entry point into the study was at the start of ART and the end-point was at the collection of an HIV VL that had a result. We present the time to first unsuppressed VL (>1000 copies/mL) as rate per 1000 person-years; time zero of this analysis was the start of ART. We set the time to elevated HIV VL data using patient identifiers in order to allow for multiple observations and multiple HIV VL results above 1000 copies per patient. Of the 1,927,570 records of HIV VL results, we analysed 893,619 (46%) complete cases with the rest being left out because they had missing dates of staring ART or dates of sample collection, processing and returning of the results to the health facility. The missing data was mainly due to the dates not consistently being filled in the hard copy laboratory request forms by the service providers ordering HIV VL samples across the various health facilities in Malawi.

### Data Analysis

A descriptive analysis was first performed detailing the characteristics of the study population. We first fitted a bivariate analysis of each of the independent variables and incidence of unsuppressed HIV VL. Only the independent variables that were statistically significant at 20% level were eligible for inclusion in the multivariable Cox model. We fitted a multivariable Cox regression model of HIV VL failure using a forward step-wise selection method, with categorized age group at VL sample and sex entered as a priori variables. We presented both crude and adjusted hazard ratios (HR) of HIV VL rebound for all included independent variables. Statistical significance was set at P<0.05.

### Ethics approval

The study was approved by the Malawi National Health Sciences Research Committee (NHSRC) in Lilongwe, Malawi (protocol#:1669). As this study used secondary anonymized data, the issue of informed consent did not apply.

## RESULTS

### Characteristics of the study participants

We evaluated 230,465 adult and children receiving ART and having a viral load test between 2011 and 2020. Of these, 63% were females; 7% were aged below 15 years while 93% were aged 15 and above, 64% were from rural areas; 10% from the northern region and 48% from the southern region; and 1% accessed HIV care at health facilities managed by private companies while 75% accessed HIV care from public facilities (Table 1). Overall, 15% of the VL sample were plasma while 85% were DBS VL samples; and 3% of the VL samples were targeted (to confirm suspected treatment failure) and 97% were taken during routine scheduled monitoring visiting. The median age at VL sample draw was 37 years (inter-quartile range (IQR): 29-45). There was an increasing trend in the number of ART patients by age (Table 1). The highest number of samples were drawn in 2019 (36%) while the least were drawn in 2011 (<1%).

**Table 1:** Characteristics of patients receiving antiretroviral therapy who had at least one HIV viral load test between 2011 and 2020 in Malawi.

| **Patient characteristics** | **n (%)** |
| --- | --- |
| **Total** | 230465 (100.0) |
| **Sex** |  |
| *Male* | 84275 (36.6) |
| *Female* | 146190 (63.4) |
| **Location** |  |
| *Rural* | 148143 (64.3) |
| *Urban* | 82322 (35.7) |
| **Age at first sample draw (in years)** |  |
| *0-4* | 3820 (1.7) |
| *5-9* | 5710 (2.5) |
| *10-14* | 6684 (2.9) |
| *15-19* | 7160 (3.1) |
| *20-24* | 14705 (6.4) |
| *25-29* | 22951 (10.0) |
| *30-34* | 34284 (14.9) |
| *35+* | 135140 (58.6) |
| *Missing* | 11 (<0.1) |
| **Region** |  |
| *Northern* | 23565 (10.2) |
| *Central* | 96979 (42.1) |
| *Southern* | 109921 (47.7) |
| **Managing authority of health facility** |  |
| *Faith-based* | 45607 (19.8) |
| *Company* | 2221 (1.0) |
| *Government* | 172497 (74.9) |
| *Private* | 10140 (4.4) |
| **Type of sample** |  |
| *DBS* | 196128 (85.1) |
| *Plasma* | 34337 (14.9) |
| **Reason for viral load sample** |  |
| *Targeted* | 6777 (2.9) |
| *Routine* | 223688 (97.1) |

DBS=Dried blood spot

### Incidence rate of HIV viral load rebound in Malawi

The median time from start of ART to having the HIV VL result was 2.9 years (interquartile range (IQR): 0.9-6.2). The incidence rates of HIV VL rebound in Malawi between 2011 and 2020 are shown in Table 2. The overall incidence rate of HIV VL rebound in Malawi between 2011 and 2020 was 16.34 (95%CI: 16.20-16.49) per 1000 person years on ART. The incidence rates of HIV VL rebound varied by age, sex, rural/urban location, region, managing authority of the health facility, year the sample was taken and reason for VL testing. As shown in Table 2, males had higher incidence of HIV VL rebound (20.17 vs 14.46, P<0.001); fewer urban than rural residents had HIV VL rebound (15.82 vs 16.66 respectively, P<0.001). There was a decreasing trend in HIV VL rebound by increasing age at HIV VL sample as well as by years on ART. The northern region had the lowest incidence of HIV VL rebound while the southern region had the highest incidence of HIV VL rebound (see Table 2). The patients that accessed HIV care from the private clinics had higher incidence of HIV VL rebound than those who accessed HIV care from company health services provided by employer or public healthcare facilities.

**Table 2:** Factors associated with the rates and rate ratios of unsuppressed HIV VL amongst the patients that were on antiretroviral therapy and that had a HIV viral load test between 2011 and 2020 in Malawi.

| Patient characteristics | Total months of follow-up | Number of patients with HIV VL >=1000 | Rate of VL >= 1000 per 1000 patient years on ART (95%CI) | Likelihood Ratio P-value | Crude RR (95%CI) | P-value | Adjusted RR (95%CI) | P-value |
| --- | --- | --- | --- | --- | --- | --- | --- | --- |
| **Total** | 2800.0 | 46084 | 16.34 (16.20-16.49) |  |  |  |  |  |
| **Gender** |  |  |  | <0.001 |  |  |  |  |
| *Male* | 931.4 | 18788 | 20.17 (19.89-20.46) |  | 1.00 |  | 1.00 |  |
| *Female* | 1900.0 | 27296 | 14.46 (14.29-14.63) |  | 0.71 (0.70-0.73) | <0.001 | 0.79 (0.77-0.80) | <0.001 |
| **Location** |  |  |  | <0.001 |  |  |  |  |
| *Rural* | 1800.0 | 29327 | 16.66 (16.47-16.85) |  | 1.00 |  | 1.00 |  |
| *Urban* | 1100.0 | 16757 | 15.82 (15.59-16.01) |  | 0.91 (0.89-0.92) | <0.001 | 0.95 (0.94-0.98) | <0.001 |
| **Age at sample draw (in years)** |  |  |  | <0.001 |  |  |  |  |
| *0-4* | 15.1 | 3062 | 202.69 (195.64-210.00) |  | 1.00 |  | 1.00 |  |
| *5-9* | 56.7 | 4625 | 81.64 (79.32-84.03) |  | 0.37 (0.36-9.39) | <0.001 | 0.39 (0.37-0.40) | <0.001 |
| *10-14* | 91.6 | 4983 | 54.38 (52.89-55.91) |  | 0.22 (0.21-0.23) | <0.001 | 0.23 (0.22-0.24) | <0.001 |
| *15-19* | 78.1 | 3290 | 42.10 (40.69-43.57) |  | 0.16 (0.15-0.17) | <0.001 | 0.18 (0.17-0.18) | <0.001 |
| *20-24* | 83.0 | 2607 | 31.42 (30.24-32.65) |  | 0.13 (0.13-0.14) | <0.001 | 0.16 (0.15-0.17) | <0.001 |
| *25-29* | 143.6 | 3399 | 23.68 (22.89-24.49) |  | 0.11 (0.10-0.11) | <0.001 | 0.14 (0.13-0.14) | <0.001 |
| *30-34* | 278.6 | 4997 | 17.94 (17.45-18.44) |  | 0.08 (0.07-0.08) | <0.001 | 0.10 (0.09-0.10) | <0.001 |
| *35+* | 2100.0 | 19121 | 9.22 (9.09-9.36) |  | 0.03 (0.03-0.04) | <0.001 | 0.04 (0.04-0.05) | <0.001 |
| **Region** |  |  |  | <0.001 |  |  |  |  |
| *Northern* | 367.2 | 5728 | 15.60 (15.20-16.01) |  | 1.00 |  | 1.00 |  |
| *Central* | 972.6 | 15762 | 16.21 (15.95-16.46) |  | 1.06 (1.03-1.09) | <0.001 | 1.04 (1.01-1.08) | 0.01 |
| *Southern* | 1500.0 | 24594 | 16.62 (16.41-16.83) |  | 1.11 (1.08-1.14) | <0.001 | 1.13 (1.10-1.16) | <0.001 |
| **Year Sample drawn** |  |  |  | <0.001 |  |  |  |  |
| *2011* | 0.4 | 19 | 52.43 (33.44-82.19) |  | 1.00 |  | 1.00 |  |
| *2012* | 6.9 | 279 | 40.70 (36.19-45.77) |  | 0.79 (0.50-1.26) | 0.32 | 0.80 (0.50-1.27) | 0.35 |
| *2013* | 14.2 | 471 | 33.06 (30.21-36.19) |  | 0.62 (0.39-0.98) | 0.04 | 0.62 (0.39-0.98) | 0.04 |
| *2014* | 49.2 | 1889 | 38.41 (36.72-40.18) |  | 0.70 (0.44-1.10) | 0.12 | 0.77 (0.49-1.21) | 0.26 |
| *2015* | 55.1 | 1977 | 35.90 (34.35-37.52) |  | 0.64 (0.41-1.01) | 0.05 | 0.70 (0.44-1.10) | 0.12 |
| *2016* | 44.4 | 1784 | 40.22 (38.40-42.13) |  | 0.71 (0.45-1.12) | 0.14 | 0.74 (0.47-1.16) | 0.19 |
| *2017* | 113.5 | 3865 | 34.04 (32.99-35.13) |  | 0.59 (0.38-0.93) | 0.02 | 0.63 (0.40-0.99) | 0.04 |
| *2018* | 273.5 | 7229 | 26.43 (25.83-27.05) |  | 0.45 (0.29-0.71) | 0.001 | 0.47 (0.30-0.74) | 0.001 |
| *2019* | 1400.0 | 16545 | 11.87 (11.69-12.06) |  | 0.19 (0.12-0.30) | <0.001 | 0.21 (0.13-0.33) | <0.001 |
| *2020* | 869.1 | 12026 | 13.84 (13.59-14.09) |  | 0.22 (0.14-0.34) | <0.001 | 0.21 (0.13-0.32) | <0.001 |
| **Managing authority of facility** |  |  |  | <0.001 |  |  |  |  |
| *Faith-based* | 577.5 | 8879 | 15.38 (15.06-15.70) |  | 1.00 |  | 1.00 |  |
| *Company* | 24.1 | 371 | 15.42 (13.93-17.07) |  | 1.04 (0.94-1.16) | 0.42 | 1.17 (1.05-1.30) | 0.004 |
| *Government* | 2100.0 | 34877 | 16.42 (16.25-16.59) |  | 1.07 (1.04-1.09) | <0.001 | 1.12 (1.09-1.15) | <0.001 |
| *Private* | 93.9 | 1957 | 20.83 (19.93-21.78) |  | 1.34 (1.27-1.40) | <0.001 | 0.71 (0.68-0.75) | <0.001 |
| **Type of sample** |  |  |  | <0.001 |  |  |  |  |
| *DBS* | 2500.0 | 39594 | 16.07 (15.91-16.22) |  | 1.00 |  |  |  |
| *Plasma* | 355.0 | 6490 | 18.28 (17.84-18.73) |  | 1.13 (1.10-1.16) | <0.001 |  |  |
| **Reason for viral load sample** |  |  |  | <0.001 |  |  |  |  |
| *Targeted* | 574.5 | 14367 | 25.01 (24.60-25.42) |  | 1.00 |  | 1.00 |  |
| *Routine* | 2200.0 | 31717 | 14.13 (13.97-14.28) |  | 0.57 (0.56-0.59) | <0.001 | 0.57 (0.56-0.58) | <0.001 |

DBS=Dried blood spot

### Trends in HIV viral load rebound amongst adults and children in Malawi

#### Temporal trends

The temporal trends of the incidence of HIV VL rebound amongst patients receiving ART in Malawi between 2011 and 2020 are shown in Figure 1. We observed a generally decreasing trend in the incidence rate of HIV VL rebound in all age and sex groups. Males had higher incidence rates of HIV VL rebound than females (Figure 1). Overall, those aged below 20 years had higher incidence rate of HIV VL rebound than the ones aged 20 years or above (P<0.001). Amongst the patients receiving ART and aged below 20 years, males had higher incidence rate of HIV VL rebound than the females (P<0.001). There incidence rate of HIV VL rebound was similar between men and women aged 20 or above. Between 2016 and 2018, there was a slight increase in the incidence of unsuppressed HIV VL and a drop thereafter. The increase was more pronounced amongst those aged below 20 years than those aged 20+ years.

**Figure 1:**
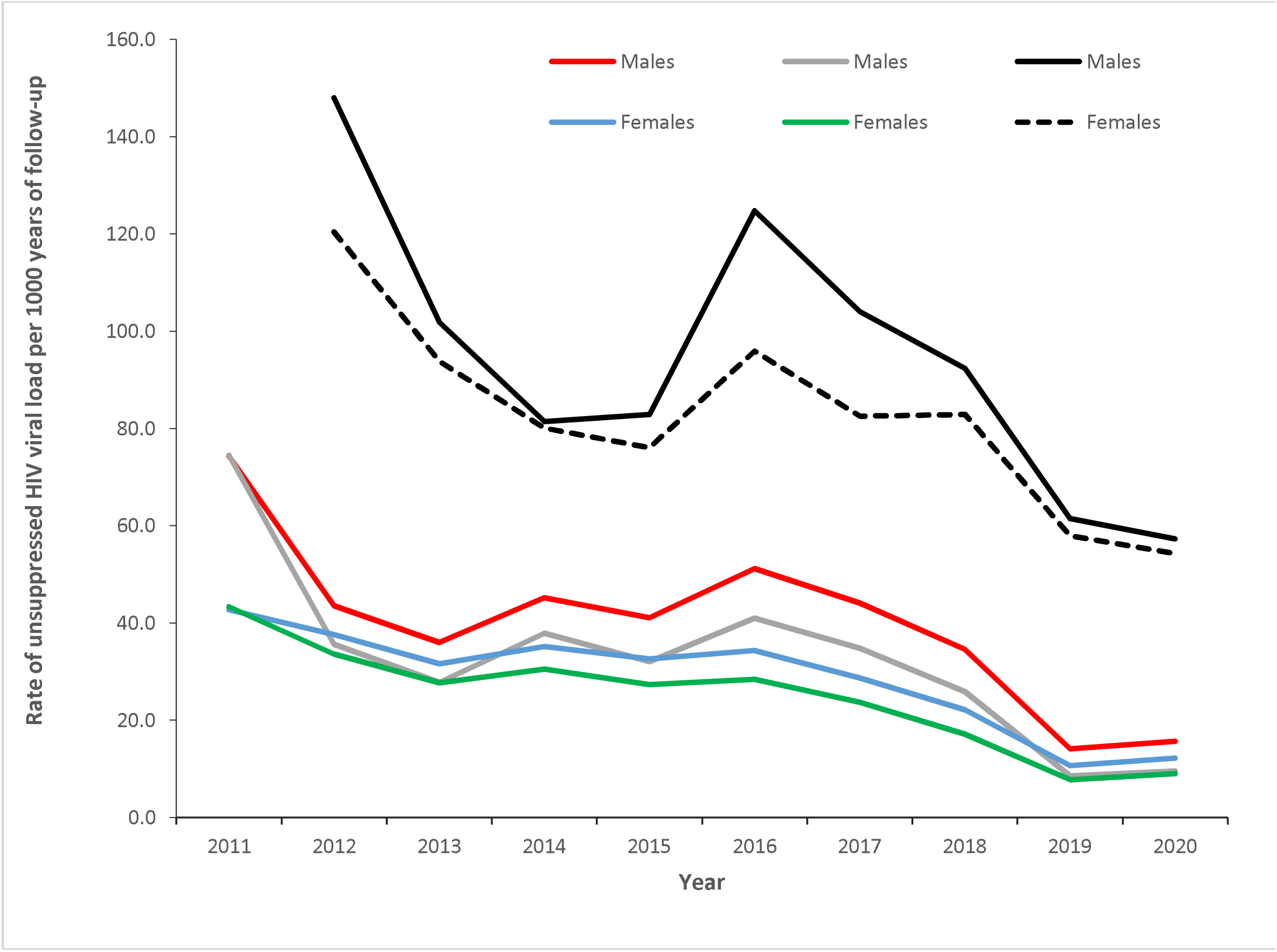
Annual trends in rates of unsuppressed HIV viral load by age and sex in Malawi between 2011 and 2020. HIV=Human Immunodeficiency Syndrome

#### Age and sex trends by months on antiretroviral therapy

The age and sex trends in the incidence rate of HIV VL rebound by months that the patients have been on ART are shown in Figure 2. As time on ART increases there is a decreasing trend in the incidence rate of HIV VL rebound.

**Figure 2:**
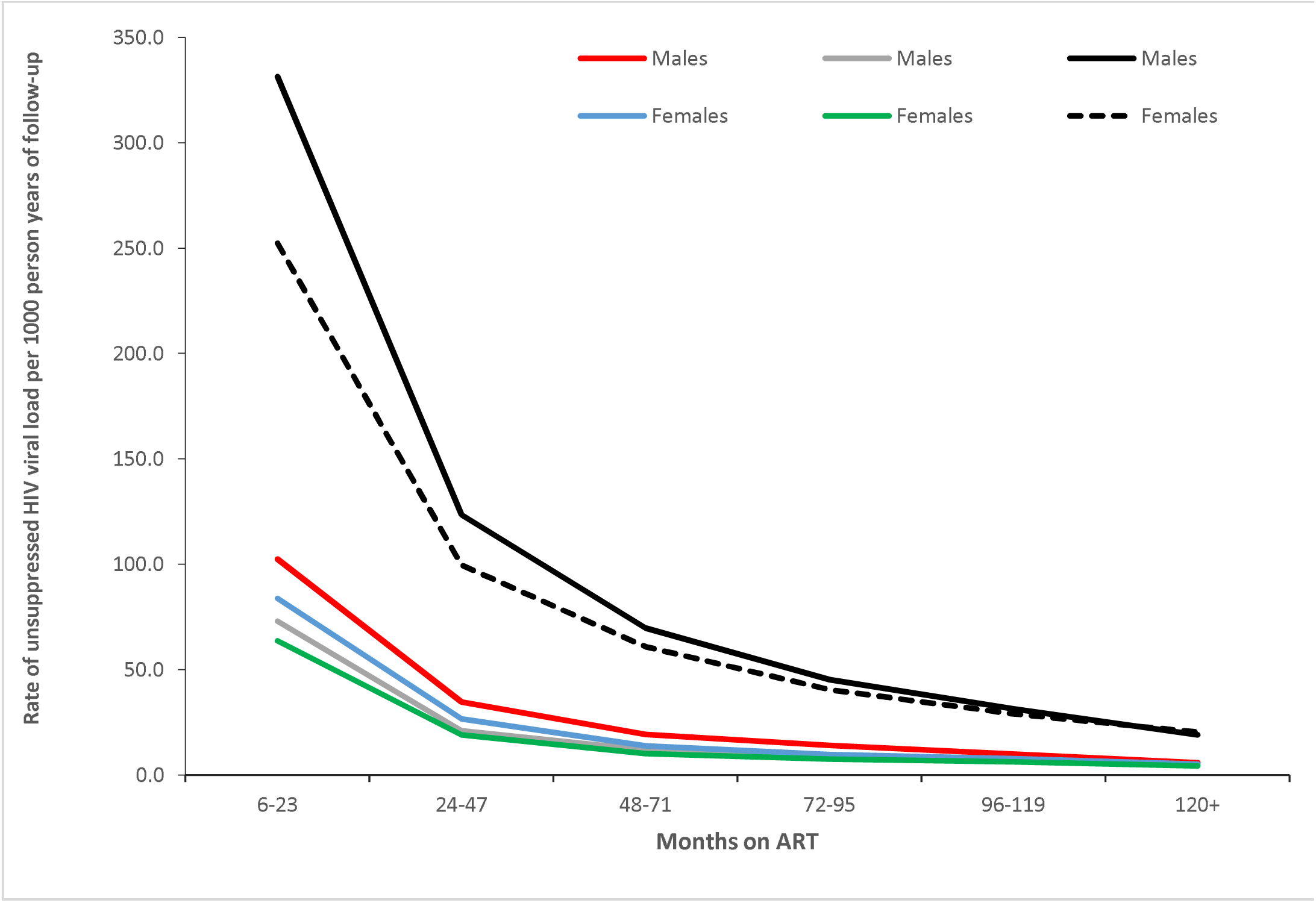
Age and sex trends in rates of unsuppressed HIV viral load by months on antiretroviral therapy in Malawi between 2011 and 2020. ART=Antiretroviral therapy

### Factors Associated with HIV Viral Load rebound in Malawi

The factors associated with the rate ratios of HIV VL rebound are shown in Table 2. After multivariable Cox regression modelling, sex, age at sample collection, year of sample collection, reason for VL sample collection, reason for collecting HIV VL sample, managing authority of the health facility, rural/urban location, and region were statistically significantly associated with the incidence of HIV VL rebound amongst Malawians that were on ART. The adjusted hazard ratio (aHR) for having HIV VL rebound amongst the female was 0.79 (95%CI:0.77-0.80, P<0.001) compared with males. The aHR of having HIV VL rebound was lower in urban than rural areas (aHR=0.95, 95% CI: 0.94-0.98, P<0.001). The aHR for having HIV VL rebound amongst the patients with routine HIV VL monitoring was 0.57 (95% CI:0.56-0.58, P<0.001) times those of the patients with targeted HIV VL monitoring. The aHR for HIV VL rebound was higher in government/public or company healthcare facilities than in faith-based health facilities (see Table 2). We observed a statistically significant decreasing trend in the hazard ratios of patients having HIV VL rebound with increasing age (P<0.001; see Table 2). The aHR for the effect of sex, rural/urban location and age at collection of the HIV VL sample were higher compared to the crude hazard ratios. However, the aHR and cHR for the reason for collecting the HIV VL were the same.

## DISCUSSION

This is the first national analysis of data for the HIV VL data from the LIMS covering all health facilities providing antiretroviral treatment in Malawi between 2011 and 2020. The following were the key findings of the study: (a) the overall incidence rate of HIV VL rebound was 16.34 (95%CI: 16.20-16.49) per 1000 person years on ART, (b) female sex, older age, rural residency, and residing in the Southern region were associated with reduction in the rate of HIV VL rebound; and (c) the rate of HIV VL rebound increased between 2012 and 2019 while the rate of HIV VL rebound reduced between 2019 and 2020.

The rate of HIV VL rebound reduced with the age of the person. This finding is consistent with other studies conducted in the Republic of South Africa (RSA) [10]; and Eswatini, Lesotho, Malawi, Zambia and Zimbabwe [7] [11] as well as Uganda [12] that found that the younger persons were more likely to HIV VL rebound than the older persons. Some of the possible explanations for higher HIV VL amongst children include non-disclosure of HIV status to the children; children being in boarding schools and not knowing why they have to take antiretrovirals every day; bitter taste of antiretrovirals taken by children; high numbers of pills; increased burden of taking other drugs; and malnutrition and lack of food negatively affecting children’s drug adherence[13]. Factors related to the health system that may explain the high rates of HIV VL rebound; distances to health facilities; and child unfriendly services [13].

Therefore, this implies the need to intensify ART adherence for paediatric HIV patients, provide antiretroviral regimens that taste sweeter and enhance more child-friendly ART clinics. In addition, the association between age and rates of suppression of HIV VL is confounded by regimen in the later period on ART. This is consistent with the general pattern of improvement in ART adherence with longer duration on ART. Therefore, the later ages ended up having higher HIV VL suppression rates.

Our findings also demonstrate differences in rates of HIV VL rebound between rural and urban areas, people in rural regions having lower rates of HIV VL rebound than the urban population. This is in consistent to what was observed in RSA [10] and Ethiopia [14] in which the urban population receiving ART had a higher likelihood of HIV VL rebound than the rural population. Some high-income countries like the USA demonstrate no statistically significant rural-urban differences in rates of HIV VL rebound [15]. The difference observed in SSA could be due to differences in the HIV/AIDS health system designs in these settings. In Malawi, HIV prevalence is much higher in urban than in rural areas as reported in the Malawi Population-based HIV Impact Assessment (MPHIA). The inequality in rates of HIV VL rebound across the rural-urban division may indicate the potential to have more resistant HIV strains in urban areas than in the rural areas [16]. Other explanations for the higher rates of HIV VL rebound in urban areas may include high in-migration that is coupled with treatment interruptions or lack of transport to the clinic for patients [17] as well as patients being busier with economic activities for their livelihood in urban areas hence skipping their ART appointments and consequently having higher rates of HIV VL rebound amongst the urban population. Therefore, introducing longer opening of clinics in urban areas as well as introduction of care groups for ART pick-up may improve ART adherence and consequently reduce rates of HIV VL rebound.

We also found that females were less likely to have HIV VL rebound than the males as reported in RSA[10]; Ethiopia[14] and Eswatini, Lesotho, Malawi, Zambia and Zimbabwe [7]. This may be due to that fact that men interrupted treatment more often than females [18]. Therefore, introduction of ART clinics with prolonged opening hours may help cover this gap by ensuring that most of the men have access to antiretrovirals and hence having suppressed HIV VL. In addition, the women tend to start ART earlier than males hence the higher likelihood to have lower rates of HIV VL rebound. Further, conducting assessment on acceptability of ART services to be provided outside the usual opening hours could be done before their actual implementation in order to focus on their acceptability by the male population.

Consistent with studies conducted in RSA[10]; Ethiopia[14] and Eswatini, Lesotho, Malawi, Zambia and Zimbabwe [7] [11] we also found that the rates of HIV VL rebound increased with duration on ART. This has practical implications on introduction of more enhanced and targeted ART adherence counselling, especially amongst persons who have just started ART. Other studies have quoted the presence of antiretroviral-related side-effects as being the key barrier to unsuppressed HIV VL amongst the persons on ART.

This study found that the southern region of Malawi had the highest rates of HIV VL rebound. Similar studies conducted in Zambia [19] and Kenya [20] have also shown regional heterogeneity in rates of HIV VL rebound. Therefore, national HIV programmes should investigate the reasons for the regional variations in rates of HIV VL rebound in order to minimize the rate of HIV VL rebound throughout the country.

The findings of this study are consistent with a study conducted in RSA which also shows an increasing trend in rates that HIV VL rebound between 2012 and 2016 [21]. Understanding the temporal trends has significant implications for improving the quality of HIV VL services provided to the patients especially if the trends are monitored and explanations for any observed change is recorded. This applies to the entire HIV programme in Malawi and other similar settings. The slight increase in the rates of the patients with suppressed HIV VL between 2016 to 2019 and a drop thereafter is linked to the most likely explained by regimen transitions in Malawi where most of the patients on first-line ART were being put on Tenofovir, Lamivudine, and Dolutegravir (TLD) from 2019. The Malawi Ministry of Health transitioned from biannual HIV VL monitoring to annual HIV VL monitoring in April 2019.

The limitations of this study included missing data on antiretroviral regimens, TB status, dates of staring ART or dates of sample collection, processing and returning of the results to the health facility. This meant that some of the analyses that have been reported by other studies could not be conducted. For example we could not assess the effect of ARV regimens or TB status [10] [22] on rate of unsuppressed HIV VL. Furthermore, absence of the data on ARV regimens taken may have resulted in spurious association between age and rates of HIV VL suppression. The other limitation of including both routine monitoring and targeted / FUP samples and patients could not be reliably re-identified. This limits the generalizability and ability to extrapolate the findings to the whole ART programme. Furthermore, it is difficult to interpret routine scheduled and targeted follow-up HIV VL results all in the same analysis, particularly since we could not reliably identify subsequent samples from the same patients from the Malawi LIMS. Due to the protocol that someone with high HIV VL on a routine monitoring sample is supposed to get intensive adherence counselling and a follow-up sample, there is a high probability that patients with a high initial VL are represented with two or more subsequent high results. It is therefore challenging to interpret and extrapolate from this since it was difficult to link some of the HIV VL results for some patients since the ART identification information were not captured consistently into the LIMS.

The strength of this study is that the sample size was big and covered the whole country of Malawi over an extensive time period. Furthermore, routine programme HIV data was used which increases the potential of the findings to inform the HIV programmes on reducing the rates of HIV VL rebound in Malawi and other similar settings. The study also has implications on improving the documentation especially for the inclusion of key variables like ARV regimens in laboratory data on HIV VL.

## CONCLUSIONS

This is the first national analysis of HIV VL data from the laboratory database management system covering all the health facilities providing antiretroviral treatment in Malawi between 2011 and 2020. Our findings show the need to take into account the geographic and demographic characteristics of the patients in order to persistently suppress HIV VL and consequently achieve the goal of achieving HIV VL suppression by 2030.

## Data Availability

Data will be available upon request from the corresponding author

## DECLARATIONS

We declare that there is no conflict of interest in publishing this paper.

## AUTHORS’ CONTRIBUTIONS

WN led the manuscript writing, conducted data management and analysis; JE advised on data analysis, AJ advised on data analysis and policy implications of the results, EE advised on data analysis, TC advised on data analysis, RN advised on policy implications of the results and OK advised on the analysis. All authors read and approved the final manuscript.

## ACKNOWLEDGEMENTS

The authors would like to thank the Malawi Ministry of Health for allowing us to use the LIMS HIV VL. The study was supported by a grant from the Swiss National Science Foundation (no 163878).

## REFERENCES

1. E. Zaniewski et al., Trends in CD4 and viral load testing 2005 to 2018: multi-cohort study of people living with HIV in Southern Africa, J. Int. AIDS Soc., vol. 23, no. 7, pp. 1–10, 2020, doi: 10.1002/jia2.25546.

2. WHO, Guideline on when to start antiretroviral therapy and on pre-exposure prophylaxis for HIV, WHO, 2016.

3. WHO, Guidelines for managing advanced HIV disease and rapid initiation of antiretroviral therapy, vol. 2. 2017.

4. World Health Organization, 2018 Global Reference List of 100 Core Health Indicators (plus health-related SDGs), WHO, 2018, [Online]. Available: https://apps.who.int/iris/bitstream/handle/10665/259951/WHO-HIS-IER-GPM-2018.1-eng.pdf?sequence=1.

5. W. Ng’ambi, I. K. Chiumia, N. Chagoma, and J. Mfutso-bengo, Factors Associated with Uptake of HIV Testing in Malawi□: A Trend Analysis of the Malawi Demographic and Health Survey Data from 2004 to 2016, J. HIV AIDS Res., vol. 2, no. 1, pp. 1–10, 2020.

6. L. Bulage et al., Factors Associated with Virological Non-suppression among HIV-Positive Patients on Antiretroviral Therapy in Uganda, August 2014-July 2015, BMC Infect. Dis., vol. 17, no. 1, pp. 1–11, 2017, doi: 10.1186/s12879-017-2428-3.

7. A. D. Haas et al., Prevalence of nonsuppressed viral load and associated factors among HIV-positive adults receiving antiretroviral therapy in Eswatini, Lesotho, Malawi, Zambia and Zimbabwe (2015 to 2017): results from population-based nationally representative surveys, J. Int. AIDS Soc., vol. 23, no. 11, pp. 1–12, 2020, doi: 10.1002/jia2.25631.

8. MoH, Department of HIV/AIDS Management Information System. Ministry of Health, Lilongwe, Malawi; 2021.

9. S. Nicholas et al., Point-of-care viral load monitoring□: outcomes from a decentralized HIV programme in Malawi, pp. 1–9, 2019, doi: 10.1002/jia2.25387.

10. Joseph Davey D, Abrahams Z, Feinberg M, et al. Factors associated with recent unsuppressed viral load in HIV-1-infected patients in care on first-line antiretroviral therapy in South Africa. International Journal of STD & AIDS. 2018;29(6):603–610. doi:10.1177/0956462417748859.

11. T. Apollo, K. C. Takarinda, A. Phillips, C. Ndhlovu, and F. M. Cowan, Provision of HIV viral load testing services in Zimbabwe: Secondary data analyses using data from health facilities using the electronic Patient Monitoring System, PLoS One, vol. 16, no. 1, p. e0245720, Jan. 2021, [Online]. Available: https://doi.org/10.1371/journal.pone.0245720.

12. L. Bulage et al., Factors Associated with Virological Non-suppression among HIV-Positive Patients on Antiretroviral Therapy in Uganda, August 2014–July 2015, BMC Infect. Dis., vol. 17, no. 1, p. 326, 2017, doi: 10.1186/s12879-017-2428-3.

13. Unicef, Understanding and Improving Viral Load Suppression in Children with HIV in Eastern and Southern Africa, Unicef, 2021.

14. G. Diress, S. Dagne, B. Alemnew, S. Adane, and A. Addisu, Viral Load Suppression after Enhanced Adherence Counseling and Its Predictors among High Viral Load HIV Seropositive People in North Wollo Zone Public Hospitals, Northeast Ethiopia, 2019: Retrospective Cohort Study, AIDS Res. Treat., vol. 2020, p. 8909232, 2020, doi: 10.1155/2020/8909232.

15. S. Weissman, W. A. Duffus, M. Iyer, H. Chakraborty, A. V. Samantapudi, and H. Albrecht, Rural-urban differences in HIV viral loads and progression to AIDS among new HIV cases., South. Med. J., vol. 108, no. 3, pp. 180–188, Mar. 2015, doi: 10.14423/SMJ.0000000000000255.

16. Ministry of Health, MPHIA 2015-2016 Collaborating Institutions, Malawi Ministry of Health, Lilongwe, pp. 2–70, 2017.

17. H. Tweya et al., Are They Really Lost? ‘True’ Status and Reasons for Treatment Discontinuation among HIV Infected Patients on Antiretroviral Therapy Considered Lost to Follow Up in Urban Malawi, PLoS One, vol. 8, no. 9, 2013.

18. A. D. Haas et al., Prevalence of nonsuppressed viral load and associated factors among adults receiving antiretroviral therapy in Eswatini, Lesotho, Malawi, Zambia, and Zimbabwe (2015-2017): Results from population-based nationally-representative surveys, medRxiv, 2020, doi: 10.1101/2020.07.13.20152553.

19. I. Sikazwe et al., Retention and viral suppression in a cohort of HIV patients on antiretroviral therapy in Zambia: Regionally representative estimates using a multistage-sampling-based approach, PLOS Med., vol. 16, no. 5, p. e1002811. May 2019, [Online]. Available: https://doi.org/10.1371/journal.pmed.1002811.

20. P. Cherutich et al., Detectable HIV Viral Load in Kenya: Data from a Population-Based Survey, PLoS One, vol. 11, no. 5, pp. e0154318–e0154318, May 2016, doi: 10.1371/journal.pone.0154318.

21. J. Larmarange et al., Temporal trends of population viral suppression in the context of Universal Test and Treat: the ANRS 12249 TasP trial in rural South Africa, J. Int. AIDS Soc., vol. 22, no. 10, pp. e25402–e25402, Oct. 2019, doi: 10.1002/jia2.25402.

22. M. B. Shiferaw et al., Viral suppression rate among children tested for HIV viral load at the Amhara Public Health Institute, Bahir Dar, Ethiopia, BMC Infect. Dis., vol. 19, no. 1, p. 419, 2019, doi: 10.1186/s12879-019-4058-4.

